# Genome sequencing of the first SARS-CoV-2 reported from patients with COVID-19 in Ecuador

**DOI:** 10.1101/2020.06.11.20128330

**Authors:** Sully Márquez, Belén Prado-Vivar, Juan José Guadalupe, Bernardo Gutierrez, Manuel Jibaja, Milton Tobar, Francisco Mora, Juan Gaviria, María García, Franklin Espinosa, Edison Ligña, Jorge Reyes, Verónica Barragán, Patricio Rojas-Silva, Gabriel Trueba, Michelle Grunauer, Paúl Cárdenas

## Abstract

SARS-CoV-2, the etiological agent of COVID-19 was first described in Wuhan in December 2019 and has now spread globally. Ecuador was the second country in South America to report confirmed cases. The first case reported in Quito, the capital city of Ecuador, was a tourist who came from the Netherlands and presented symptoms on March 10th, 2020 (index case). In this work we used the MinION platform (Oxford Nanopore Technologies) to sequence the metagenome of the bronchoalveolar lavage (BAL) from this case reported, and subsequently we sequenced the whole genome of the index case and other three patients using the ARTIC network protocols. Our data from the metagenomic approach confirmed the presence of SARS-CoV-2 coexisting with pathogenic bacteria suggesting coinfection. Relevant bacteria found in the BAL metagenome were *Streptococcus pneumoniae, Mycobacterium tuberculosis, Staphylococcus aureus* and *Chlamydia* spp. Lineage assignment of the four whole genomes revealed three different origins. The variant HEE-01 was imported from the Netherlands and was assigned to B lineage, HGSQ-USFQ-018, belongs to the B.1 lineage showing nine nucleotide differences with the reference strain and grouped with sequences from the United Kingdom, and HGSQ-USFQ-007 and HGSQ-USFQ-010 belong to the B lineage and grouped with sequences from Scotland. All genomes show mutations in their genomes compared to the reference strain, which could be important to understand the virulence, severity and transmissibility of the virus. Our findings also suggest that there were at least three independent introductions of SARS-CoV-2 to Ecuador.

**IMPORTANCE:** COVID-19, an infectious disease caused by SARS-CoV-2, has spread globally including Latin American countries including Ecuador. The first strain of SARS-CoV-2 sequenced was from Wuhan, which is considered as the reference strain. There were no data about the SARS-CoV-2 lineages in Ecuador, and the purpose of this study was to find out the origin of the different lineages circulating in the population. We also were interested in the mutations present in these genomes as they can influence virulence, transmission and infectivity.

## INTRODUCTION

SARS-CoV-2, the etiologic agent of COVID-19, has spread globally reaching all continents^1,2^. Ecuador was the second country in South America, after Brazil, to report its first case on 29 February 2020^3^. The number has grown since, reaching 39,098 laboratory-confirmed cases and 3,358 deaths until May 31st, 2020 ^4^. The pandemic has had a severe impact in Ecuador, as death excess estimates suggest that up to 182.44 people per million inhabitants may have died since the first COVID19 case ^5^, making Ecuador the country with the highest mortality rate in South America.

Quito, the capital city of Ecuador, was the second city to report COVID-19 cases after Guayaquil. After a severe wave of cases in Guayaquil during the early stages of the outbreak, Quito currently reports the largest number of COVID-19 daily cases in the country. The first case in Quito was diagnosed initially in the Sucumbíos province, while the patient was doing tourism, and was subsequently brought to a third-level hospital in the capital. The patient is originally from the Netherlands and was diagnosed by the Health Ministry on March 10th, 2020. His condition deteriorated while in Sucumbíos, prompting his transfer to Quito where he was admitted into the Intensive Care Unit of the Eugenio Espejo Hospital. After 32 days, the patient recovered and returned to his native country. Currently, local authorities have confirmed 3676 cases ^4^ and widespread community transmission is actively happening.

Understanding the local epidemiology of SARS-CoV-2 can greatly be enhanced from understanding the virus evolution. It is therefore relevant to invest in SARS-CoV-2 genome sequencing as it could help to establish the origin of the pathogen ^6^, its transmission across different regions ^7^, the genetic diversity of the circulating strains in the population and to identify notable mutations ^8^. Additionally, key biological aspects such as virulence, transmissibility and infectivity of the circulating strains can be investigated, when combined with clinical data from the patients ^7^. Around 35,473 genome sequences of SARS-CoV-2 have been reported from countries around the world and are available in GenBank and the GISAID (Global Initiative on Sharing All Influenza Data) repository ^9^.

In this work we describe the first four SARS-CoV-2 whole genome sequences obtained from Ecuador. We also report the metagenomic analysis of the first COVID-19 patient diagnosed in Quito. Both approaches were performed using the portable MinION platform (Oxford Nanopore Technologies).

## MATERIALS AND METHODS

### Ethics

This study used excess respiratory samples after being officially confirmed with positive COVID19 diagnosis. Ethical approval for all samples was given by CEISH-USFQ (Comité de ética de investigación en seres humanos-USFQ): IE-JP067-2020-CEISH-USFQ.

### Epidemiological information and sample collection

Samples from the four patients were collected in Hospital Eugenio Espejo (HEE) or Hospital IESS Sur (HGSQ), two public third-level hospitals at Quito, Ecuador. Sample positivity for SARS-CoV-2, using standard RT-PCR approach, was officially reported to hospitals by the Ecuadorian Ministry of Public Health (MSP) and National Institute of Public Health and Research (INSPI). Sample HEE-01 was a bronchi-alveolar lavage (BAL) from a patient of Dutch origin in his late fifties who presented symptoms during a visit to the Sucumbios province and then transferred to the HEE intensive care unit (ICU) in Quito. This patient was the first confirmed COVID-19 case in Quito. Samples HGSQ-USFQ-018, HGSQ-USFQ-007 and HGSQ-USFQ-010 were all nasopharyngeal swabs. HGSQ018 was collected from a 27 years old male patient who presented severe COVID-19 symptoms after a visit to Guayaquil and was admitted to the pneumology unit at HGSQ. Sample HGSQ-USFQ-007 was collected from a 40 years old male patient that had no history or contact with a COVID-19 patient, presented severe COVID19 symptoms and was admitted to ICU at HGSQ. Sample HGSQ-USFQ-010 was collected from a 39 years old male patient that had contact with patient HGSQ-USFQ-007 and was also admitted to ICU. The BAL sample was collected in a sterile tube with 2X DNA/RNA Shield (Zymo), and nasopharyngeal swabs were submerged in 1X DNA/RNA Shield (Zymo) to ensure virus inactivation and to preserve the genetic material. Samples were transported immediately at 4°C to the Institute of Microbiology at USFQ (IM-USFQ) in a sealed container with all the biosecurity and containment measures recommended by the CDC of the USA (https://www.fda.gov/media/134922/download).

### RNA extraction

The genetic material from samples was extracted in a biosafety type II chamber with HEPA filters in the Virology Laboratory of the Microbiology Institute (IM) at USFQ. Two different sequencing approaches were performed with the extracted RNA: metagenomics with sample HEE01 and whole genome sequencing with samples HEE01, HGSQ-USFQ-018, HGSQ-USFQ-007 and HGSQ-USFQ-010. QIAamp® Viral RNA Extraction Kit (Qiagen) was used to extract RNA from the HEE1 sample that was used for the metagenomics approach. Briefly, 250 µl of BAL sample and 560 µl of AVL lysis buffer were incubated for 1 hour, then manufacturer RNA kit instructions were followed. Sample was eluted in a final volume of 70 µl. RNA was immediately purified µsing RNA Clean and Concentrator kit (Zymo), 14 µl of purified ARN were used for retrotranscription of RNA to cDNA following the RNA Viral Metagenomics MinION One-Pot Sequencing Protocol from the genomics department of Public Health England^10,11^.

SV Total RNA Isolation System kit (Promega, USA) was used to extract RNA from samples used for the whole genome sequencing approach. A predigestion step was added to the RNA extraction protocol of the bronchoalveolar fluid (BAL) sample as follows. Before nucleic acid extraction, 280µl of the Bronchoalveolar fluid (BAL) sample was predigested with 360µl of PureLink™ Genomic Lysis Buffer and 20 µl of proteinase K. The mix was incubated at 55°C for 10 minutes, with vortexing every 5 minutes (Life Technologies, USA). All RNA extractions were performed following manufacturer instructions and eluted in a final volume of 50 µl. Total RNA was quantified using QuBit (Thermo Fisher Scientific) with a Qubit RNA Assay Kit (Thermo Scientific, Invitrogen, Carlsbad, CA, USA). Retrotranscription of RNA to cDNA was carried out using the Protocol of the Public Health England Genomics Lab ^10,11^ at the USFQ Bioinformatics Center.

### Viral whole genome sequencing approach

Primer Scheme approach developed by the ARTIC network for nCoV-2019 (Robertson, 2020) using the V1 primer sets to generate an amplicon tiling path across the viral genome ^12^. cDNA MinION library preparation was performed using the Rapid Barcoding kit (SQK-RBK004) following manufacturer instructions and then loaded into a MinION flow cell (FLO-MIN 106). Base calling of FAST5 files was performed using Guppy (version 3.4.5) (Wick, et al 2019), and the RAMPART software (v1.0.5) from the ARTIC Network (https://github.com/artic-network/rampart) was used to monitor sequencing in real-time. Sequence quality scoring, demultiplexing and adapter removal was performed with the NanoPlot ^13^ and Porechop algorithms respectively. The ARTIC Network bioinformatics pipeline was used for variant calling, and the reads were mapped against the reference strain Wuhan-Hu-1 (GenBank accession number MN908947), to generate consensus genomes. The genome was uploaded to the CoV-GLUE resource ^14^, to determine the epidemiological linkage of circulating SARS-CoV-2 variants.

### Metagenomic approach and bioinformatic analysis

cDNA MinION library preparation was performed using the Rapid Barcoding kit (SQK-RBK004) following manufacturer instructions. The resulting library was loaded on an Oxford MinION flowcell (FLO-MIN 106) and sequenced using the MinKNOW version 4.05, for 24 hours. Basecalling and quality control analysis was performed using Guppy (version 3.4.5) and NanoPlot (version 1.29.0) respectively^13^. Adapters and barcodes were removed from the MinION reads using Porechop (version 0.2.4) (https://github.com/rrwick/Porechop). Taxonomic classification of the sequences was performed using the Kaiju platform^15^. A SARSCoV-2 consensus sequence was obtained by mapping reads against the reference strain Wuhan-Hu-1 (GenBank accession number MN908947) using minimap2 (version 2.14-r883)^16^. Finally, Samtools (version 1.9) (http://samtools.github.io) and Tablet alignment viewer (version 1.19.09.3) (https://ics.hutton.ac.uk/tablet) were used to visualize the mapped sequence.

## RESULTS

### Metagenomic sequencing confirms SARS-CoV-2 infection in patient HEE-01 and possible bacterial coinfection

The metagenomic analysis found a total of 206,111 DNA sequences with 43,603,091 bases. Viral sequences represented 0.9%, of which 83% corresponded to non-assigned coronavirus and 17% were identified as SARS coronaviruses (Figure 1). The sequences assigned to coronavirus were extracted and mapped using the Wuhan-Hu-1 reference genome (GenBank accession number MN908947). A sequence similarity of 99.68% was found with this sequence with 100% query coverage. Additionally, a phylogenetic tree was generated with the sequences found and the reference strains used in GenBank NCBI. The phylogenetic alignment grouped the query sequence with the ORF1AB segment of the virus polyprotein (it encodes replication genes). The metagenome sequences are publicly available at https://www.ncbi.nlm.nih.gov/bioproject/?term=PRJNA613094 (Accession: PRJNA613094).

**Figure 1.**
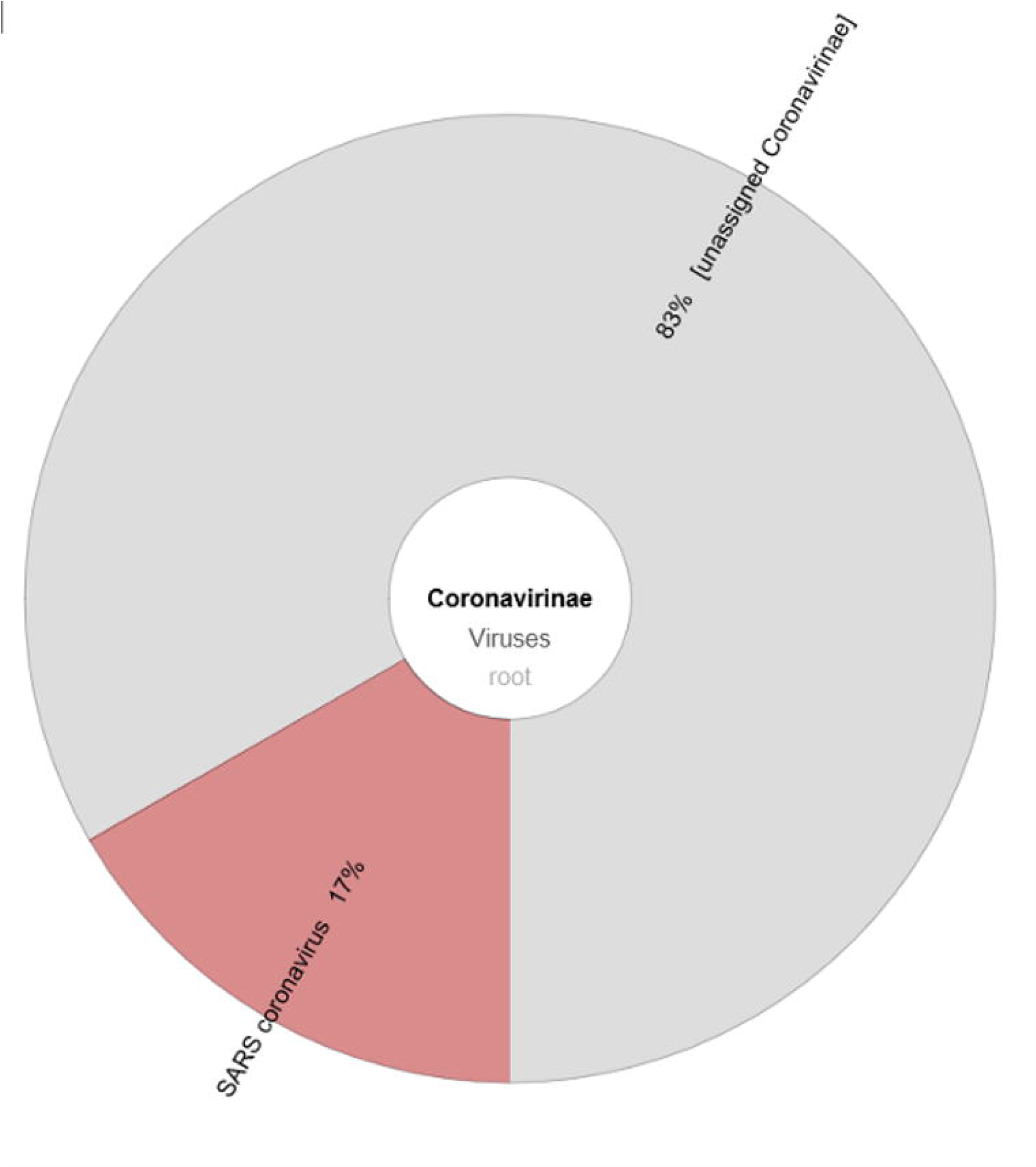
Krona chart summarizing the percentage of sequences assigned to Coronavirus in the metagenome. Coronavirus sequences represented 0.036% of all microorganisms.

Several bacterial and eukaryotic sequences related to the patient’s respiratory microbiota were identified by metagenomics. The most relevant taxa found were *Streptococcus pneumoniae, Mycobacterium tuberculosis, Staphylococcus aureus* and *Chlamydia* spp. We did not identify any particular clinically relevant fungus. To complement the metagenomic approach we carried on with the whole genome sequencing of the current sample and additionally with 3 more additional samples from severely ill patients diagnosed in March.

### Complete genome sequencing revealed multiple entries of the virus to Ecuador

Whole genome sequencing of the first case of COVID-19 in Quito revealed it to be an imported case from Europe (strain named HEE-01). Compared with the Wuhan-Hu-1 reference strain, HEE-01 shows only two nucleotide differences that include one amino acid replacement. HEE-01 was assigned to the B lineage and grouped with sequences from the Netherlands (Figure 2). Strain HGSQ-USFQ-018 was obtained from a case detected in Quito, but the infection was presumably acquired in Guayaquil. It shows nine nucleotide differences with the reference genome (Table 1). This genome is placed in the B.1 lineage and grouped with genomes from other European countries (Figure 2). Finally, HGSQ-USFQ-007, a community acquired infection in Quito, and the sequence from an epidemiologically-linked contact (HGSQ-USFQ-010) were also placed in the B lineage. Both sequences have multiple differences compared to the reference strain, including 17 amino acids replacement and one amino acid deletion (Table 1).

**Figure 2.**
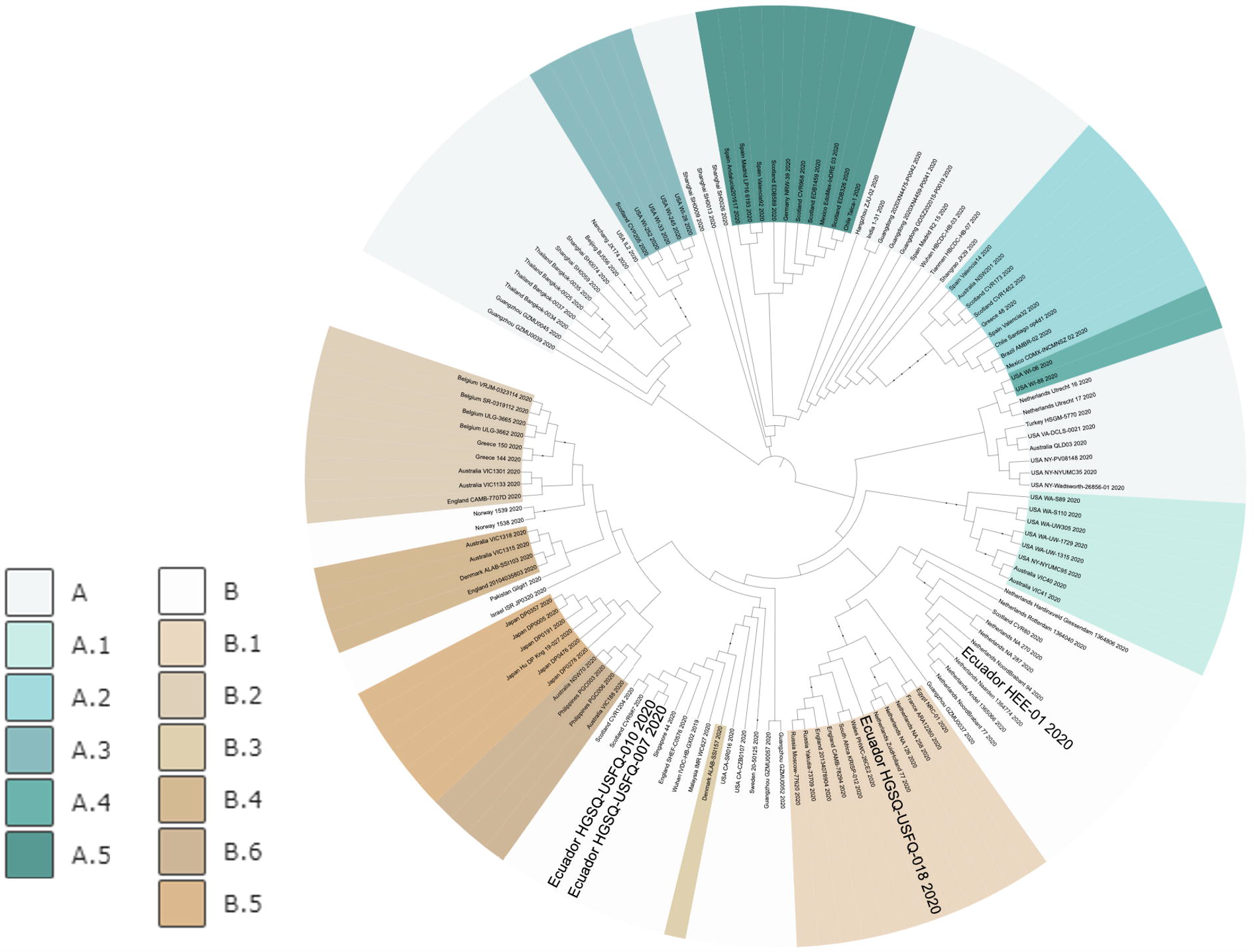
Maximum parsimony phylogenetic tree. Ecuadorian sequences are highlighted in bold. Lineage assignments are highlighted in similar colors: lineage A (Green color palette) and lineage B (Brown color palette).

**Table 1.** Mutations found in four SARS-CoV-2 Genomes from Ecuador compared to Wuhan-Hu-1 (GenBank accession number MN908947).

| Genome Sequence ID | GISAID ID | Lineage | Gene | Mutation | Amino acid | Type |
| --- | --- | --- | --- | --- | --- | --- |
| HEE-01 | EPI_ISL_417482 | B | ORF1ab | T514C | - | - |
|  |  |  | S | A25182T | E1207V | Transversions |
| HGSQ-USFQ-018 | EPI_ISL_422563 | B.1 | ORF1ab | C3037T | no | Transition |
|  |  |  | ORF1ab | C4010T | L431F | Transition |
|  |  |  | ORF1ab | C14408T | P323L | Transition |
|  |  |  | ORF1ab | C18555T | no | - |
|  |  |  | S | A23403G | D614G | Transition |
|  |  |  | ORF3a | A25983G | no | - |
|  |  |  | N | G28881A | R203K | Transition |
|  |  |  | N | G28882A | no | - |
|  |  |  | N | G28883C | G204R | Transversions |
| HGSQ-USFQ-007 | EPI_ISL_422564 | B | ORF1ab | A1996C | - | - |
| and |  |  | ORF1ab | C1997T | no | Transition |
| HGSQ-USFQ-010 | EPI_ISL_422565 | B | ORF1ab | T11737G | N255K | Transversions |
|  |  |  | ORF1ab | T20497A | N255K | Transversions |
|  |  |  | S | G22424T | A288S | Transversions |
|  |  |  | S | C24418T | no | Transition |
|  |  |  | ORF3a | A25505C | Q38P | Transversions |
|  |  |  | ORF3a | G25647T | L85F | Transversions |
|  |  |  | ORF3a | T25676C | L95S | Transition |
|  |  |  | ORF3a | C25678T | L96F | Transition |
|  |  |  | ORF3a | C25692T | no | Transition |
|  |  |  | ORF3a | T25695G | - | - |
|  |  |  | ORF3a | C25708T | L106F | Transition |
|  |  |  | ORF3a | C25710T | no | Transition |
|  |  |  | ORF3a | T25711C | Y107H | Transition |
|  |  |  | ORF3a | C25728T | no | Transition |
|  |  |  | ORF3a | A25741G | S117G | Transition |
|  |  |  | ORF3a | A25756G | R122E | Transition |
|  |  |  | ORF3a | G25757A | no | Transition |
|  |  |  | ORF3a | A25764T | - | - |
|  |  |  | ORF3a | T25779G | - | - |
|  |  |  | ORF3a | G25781A | C130* | Transition |
|  |  |  | ORF3a | C25782A | - | - |
|  |  |  | ORF3a | G25879A | V163T | Transition |
|  |  |  | ORF3a | T25880C | no | Transition |
|  |  |  | ORF3a | C25883A | T164N | Transversions |
|  |  |  | ORF3a | T25885G | S165A | Transversions |
|  |  |  | ORF8 | A28035G | R48G | Transition |
|  |  |  | ORF8 | A28037G | no | Transition |
|  |  |  | ORF8 | G28038T | V49L | Transversions |
|  |  |  | ORF8 | C28076T | no | Transition |
|  |  |  | ORF8 | T28078C | V62A | Transition |

Lineages were assigned using Pangolin COVID-19 Lineage Assigner (pangolin.cog-uk.io).

## DISCUSSION

The mitigation of the spread of SARS-CoV-2 has been unsuccessful in most South American countries. Countries like Ecuador, where the population, despite shocking examples in other cities like Guayaquil, continues to react in disbelief at an imminent risk. It is now when scientific research is needed to guide the authorities to develop correct public health policies. In this study, we were able to uncover the first 4 SARS-CoV-2 whole genome sequences from Ecuador using the portable low-cost MinION platform (Oxford Nanopore). Genome sequences, from the first COVID19 case reported in Quito, have few differences compared to the reference strain (Wuhan-Hu-1), and is grouped within lineage B near sequences from patients in the Netherlands. This finding confirms that the patient most likely acquired the infection in his country of origin. Metagenome sequencing of the patient’s BAL revealed possible bacterial coinfection as several of the microorganisms are not considered members of the core lower airways bacterial microbiota^17^. As consequence, the encountered bacteria were probably contributing to the patient’s severe condition. Metagenomic analysis could be crucial in COVID-19 cases because it adds valuable information about concurrent pulmonary pathogens that can aggravate the clinical condition^18–20^.

acquired the infection at the community level, either in Guayaquil or Quito. All three patients presented severe respiratory symptoms. Sequence recovered from a patient that got infected in Guayaquil grouped in lineage B.1, suggesting a second introduction of the virus into the country. Finally, the virus sequences recovered from two patients infected in Quito had marked differences with the previous sequences, suggesting a third introduction of the virus to the country. These sequences were identical between them, which was expected since the patients were family members, developed symptoms at the similar time and lived together. Furthermore, the distinct set of mutations from these sequences was analyzed in a separate study which reports a potential virulence factor not previously described, which is associated with inhibition of the interferon response *in vitro* ^21^. Community acquired transmission is now widespread in Ecuador and more samples from community acquired infections are needed in order to inform analyses of the local epidemiology of the virus. Comparing these genomes will allow us to provide a more accurate estimate of the arrival times and routes of the virus into certain areas, and to better understand how it is locally spreading and evolving. Revealing the information contained in the SARS-CoV-2 virus genome sequences is a robust tool to understand the epidemiology of COVID-19 locally and contribute to the global panorama.

The current pandemic is unlikely to be the last, and it is therefore essential to improve the response capacity of our public health systems and to implement and strengthen continuous scientific research programs. Only in this way we will be able to better understand these types of threats, act based on evidence, and thus reduce their impact.

## Data Availability

Sequences obtained are publicly available on GISAID under the submissions: HEE01, HGSQ-USFQ-018, HGSQ-USFQ-007 and HGSQ-USFQ-010

https://www.gisaid.org

## AKNOWLEDGEMENTS

This work was funded by Universidad San Francisco de Quito and CADDE project (www.caddecentre.org/). P.C. is funded by NIH FIC D43TW010540 Global Health Equity Scholars.

We acknowledge the submitting authors and institutions of the sequences of SARS CoV 2 from GISAID Database.

## AUTHOR CONTRIBUTIONS

Conceptualization: M.J., G.T., M.G., P.C. Methodology: S.M., B.P.-V., J.J.G., M.T., P.C. Software: B.P.-V., V.B., P.C. Validation: B.P.-V., V.B. and P.C. Formal analysis and investigation: V.B., P.R.-S. and P.C. Writing - Original Draft: P.R.-S. and P.C. Writing - Review & Editing: All authors. Visualization: V.B. and P.C. and M.A. Supervision: G.T. Funding acquisition: M.G, G.T. & P.C.

